# A Novel Hybrid LSTM-DNN Model for Ventilator Pressure Prediction: Comparative Analysis of Data Splitting Strategies

**DOI:** 10.1101/2025.03.31.25324938

**Authors:** Ali Wannous

## Abstract

In recent years, deep learning has significantly transformed ventilator pressure forecasting, which is crucial for providing patients with safe, personalized respiratory therapy. This work introduces a novel hybrid model that combines Deep Neural Networks (DNN) with Long Short-Term Memory (LSTM) networks. To enhance forecast accuracy, we incorporated a special forget gate reset mechanism to this model. Our approach, which involved meticulous data collection and analysis, produced a robust model architecture that effectively captures the unique characteristics of each breath cycle. We examined various data-splitting techniques, particularly comparing Timeseries Split and K-fold cross-validation to determine the most effective one. According to our findings, Timeseries Split performs better at preserving the sequential order of the ventilator data. Notably, compared to traditional methods, our hybrid model achieved an astounding 84% reduction in Mean Absolute Error (MAE) and a 97 % reduction in Mean Squared Error (MSE). These results highlight the potential of our approach to greatly improve ventilator control by making precise, data-driven predictions.

## Introduction

Mechanical ventilation has been a fundamental component of critical care medicine since its life-saving introduction during the poliomyelitis epidemics of the 1950s [24]. Since then, it has evolved into a vital intervention, particularly for patients with acute respiratory failure [26]. In emergency situations, pressure ventilation is crucial for addressing severe oxygen deprivation and providing critical support for patients unable to maintain independent ventilation [10] [5] [13].

Mechanical ventilation, while lifesaving, poses complications, especially in patients with lung or cardiac conditions. Accurately predicting ventilator pressure is crucial, influenced by airway compliance and resistance variability, complicating sufficient ventilation while reducing the risk of ventilator-induced lung injury [33] [4] [9].

This study introduces a hybrid LSTM-DNN model to enhance ventilator pressure prediction, leveraging LSTM for temporal dependencies and DNN for complex non-linear relationships [7] [10]. A forget gate reset mechanism minimizes variability across breath cycles, and model performance is assessed via k-fold and time-series cross-validation.

The structure of the paper is organized as follows: Section 2 reviews related studies, Section 3 discusses the theory and methodology, including data acquisition, preprocessing, and hybrid model design. Section 4 outlines the evaluation techniques along with the experimental results. Finally, Sections 5 and 6 provide the discussion and conclusion, respectively.

## Related Studies

Recent advancements in deep learning, especially hybrid models combining DNNs and LSTMs, have improved prediction tasks. Notably, AL-Ghamdi et al. [7] developed a hybrid DNN-LSTM model that significantly reduced forecasting errors in household energy consumption predictions. Building on these findings, Ozcan et al. [11] developed a hybrid DNN-LSTM model that enhanced phishing URL classification accuracy.

In healthcare, particularly mechanical ventilation, Wadne et al. [17] highlighted LSTM’s superiority in pressure prediction for lung circuits. Furthering this research, Jafril Alam et al. [10] proposed a hybrid model that utilized Bi-directional LSTM (Bi-LSTM) and Bi-directional Gated Recurrent Units (Bi-GRU) with the SELU activation function, achieving reduced error metrics and showcasing practical applications in prediction tasks. Similarly, Dief et al. [9] introduced a hybrid LSTM model enhanced by the Chimp Optimization Algorithm (ChoA), which improved hyperparameter selection and predictive accuracy. Furthermore, Abdelghani Belgaid [14] proposed a DNN approach for simulating airway pressure during the inspiratory phase, effectively tracking pressure waveforms and outperforming traditional methods. Collectively, these studies underscore the effectiveness of hybrid deep learning models and inform our implementation of a tailored DNN-LSTM approach for pressure prediction.

## Materials and methods

### Data Acquisition and Preprocessing

The ventilator data used in this study was generated with a modified open-source ventilator, developed in collaboration with Princeton University and Google Brain [14] [21]. This ventilator is connected to an artificial bellows test lung via a respiratory circuit. The datasets utilized for our research were sourced from Kaggle and are available for access at Kaggle Ventilator Pressure Prediction.

**Fig 1**. Illustration of the respiratory circuit used in the ventilator setup [Figure1]

Figure 1 shows the system configuration, including airway pressure and control inputs. Table 1 provides details on significant features, while positive end-expiratory pressure (PEEP) prevents lung collapse [9] [14].

**Table 1.** Summary of Dataset Features and Target Variable.

| Column Name | Description |
| --- | --- |
| id | Globally unique time step identifier across the entire file |
| breath_id | Globally unique time step for breaths |
| R | Lung attribute indicating how restricted the airway |
| C | Lung attribute indicating how compliant the lung |
| time_step | The actual time stamp |
| u_in | Control input for the inspiratory solenoid valve |
| u_out | Control input for the exploratory solenoid valve |
| pressure | Airway pressure measured in the respiratory circuit |

In this study, we analyzed a dataset comprising a total of 6,036,000 samples. Importantly, the dataset contains no null values, as evidenced by a 0% rate of missing entries.

Table 2 summarizes the dataset statistics, which are crucial for understanding model performance.

**Table 2.** Summary of Key Features Statistics.

| Column | Mean ± (Max – Min) · Std |
| --- | --- |
| time_step | $1.307225 \pm (2.937238 - 0) \cdot 0.7659778$ |
| u_in | $7.321615 \pm (100 - 0) \cdot 13.43470$ |
| pressure | $11.22041 \pm (50 - 5) \cdot 8.109703$ |

The figure2 illustrates the frequency of values for the discrete features.

The dataset includes 80 samples for each breath cycle, resulting in a total of 75,450 identified breath cycles in the data [9] [14].

Figure 3 illustrates the first five breath cycles, providing a visual representation of the respiratory patterns within the dataset.

**(a)** Histogram of R [Figure2]

**(b)** Histogram of C [Figure3]

**(c)** Histogram of u out [Figure4]

**Fig 2**. Histogram of discrete features [Figure2, Figure3, Figure4]

**Fig 3**. First five breath cycles illustrated [Figure5]

**(a)** Mean of Airway Pressure [Figure6] **(b)** Variance of Airway Pressure [Figure7]

**Fig 4**. Mean and Variance of Airway Pressure [Figure6, Figure7]

Calculating the mean and standard deviation across breath cycles, as shown in Figures 4, reveals significant trends: mean values rise to about 20 in the first 40 samples, then drop to 6, with variance stabilizing, indicating consistent pressure maintenance.

1. Magnitude [Figure8] **(b)** Phase [Figure9]

**Fig 5**. Magnitude and Phase of Fourier coefficients for mean airway pressure [Figure8, Figure9]

Figures 5 show that the Fourier coefficients for mean pressure remain zero across breath cycles, except at the initial and final points, highlighting pressure behavior over time.

**Fig 6**. Pearson correlations for features [Figure10]

Figure 6 illustrates a low overall correlation of 0 between pressure and time, but within individual breath cycles, a notable negative correlation of −0.71 emerges, underscoring the importance of considering temporal context for effective pressure prediction.

This comprehensive data analysis and preprocessing are essential for creating a robust hybrid LSTM-DNN model that can accurately predict airway pressure, leading to improved patient outcomes.

### Design of the Hybrid Model

The hybrid model combines the strengths of Long Short-Term Memory (LSTM) networks and Deep Neural Networks (DNNs). LSTMs are excellent at processing sequential data, while DNNs are powerful for extracting complex features [7] [10]. Together, this hybrid model is designed to effectively capture both the time-dependent patterns and non-linear relationships that are crucial for predicting ventilator pressure.

#### DNN Component

Deep Neural Networks (DNNs) represent a significant advancement in artificial intelligence and machine learning, particularly for modeling complex, non-linear relationships in data. Composed of multiple layers of interconnected neurons, DNNs transform input data into output [25] [7]. Each neuron processes inputs through a linear transformation with learned weights and biases, followed by an activation function that introduces non-linearity, enabling the capture of intricate patterns [7]. This multi-layered architecture allows DNNs to learn hierarchical data representations, making them effective for various applications in both supervised and unsupervised learning contexts [31].

##### Definition

Let 𝒩 be a neural network defined as follows:

1. Let *d* ∈ ℕ represent the dimension of the input layer.
2. Let *L* denote the total number of layers in the network.
3. Let *N*_*l*_ denote the number of neurons in layer *l*, where *l* = 0, 1, …, *L*. Specifically, *N*_0_ = *d* represents the quantity of neurons in the input layer.
4. Let 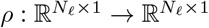 be a non-linear activation function applied at each layer, defined as:

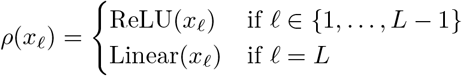

The neural network 𝒩 transforms an input vector *x* ∈ ℝ^*d*^ to the real number space ℝ through the following transformations:

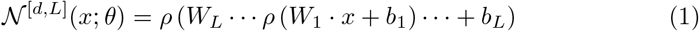

where 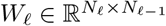 is the weight matrix associated with layer 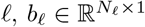 is the bias vector for layer *l*, and *θ* includes all the parameters of the network, such as the weights and biases for each layer.

This formal definition captures the structure and functioning of a neural network, emphasizing the role of activation functions and the relationship between input, hidden, and output layers.

#### LSTM Component

Long Short-Term Memory (LSTM) networks represent a distinctive type of Recurrent Neural Network (RNN) designed specifically to capture and model long-term dependencies in sequential data [10] [3]. Equipped with a memory cell and a gating mechanism, LSTMs can selectively remember or forget information, overcoming the limitations of traditional RNNs that struggle with long sequences [7]. This architecture, featuring forget, input, and output gates, enables LSTMs to efficiently handle long-range temporal dependencies, making them especially well-suited for time-series data like ventilator pressure readings [2] [3]. The cell state facilitates the preservation of information flow throughout the sequence, as illustrated in Figure 7 [27].

**Fig 7**. The structure of LSTM cell [Figure11]

1. The Forget Gate *f*_*t*_ The forget gate *f*_*t*_ determines which information is retained from the previous cell state [23] [7]. It processes the current input *x*_*t*_ and previous hidden state *h*_*t*−1_ using weights *W*_*f*_ and bias *b*_*f*_, passed through a sigmoid function [19] [23] [7]:

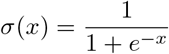

The forget gate is computed as:

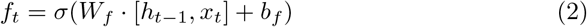

This mechanism enables LSTMs to learn long-term dependencies effectively [3].
2. The Input Gate *i*_*t*_ The input gate *i*_*t*_ determines the amount of new information is stored in the cell state [23] [7]. It computes a candidate cell state 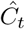 from the previous hidden state *h*_*t*−1_ and current input *x*_*t*_ using weights *W*_*c*_ and bias *b*_*c*_ [6]:

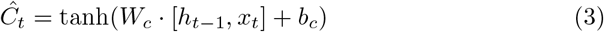

The input gate then regulates the candidate state with:

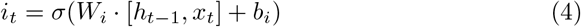

This mechanism enables efficient learning by controlling the retention of new information [23] [7].
3. The Output Gate *o*_*t*_ The output gate *o*_*t*_ controls the transfer of information from the cell state to the hidden state, deciding which elements of the cell state contribute to the calculation of the hidden state [23] [7]. The cell state *C*_*t*_ is updated as follows:

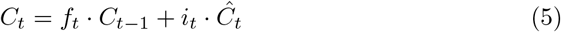

The output gate activation is computed by:

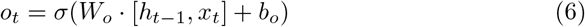

The hidden state *h*_*t*_ is then given by:

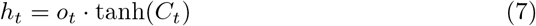

This mechanism ensures effective modeling of long-term dependencies [23] [7].

#### Hybrid Model Design

The proposed hybrid model integrates two parallel Deep Neural Networks (DNN1 and DNN2), which operate in tandem with an LSTM network to capture both temporal dependencies and non-linear relationships in the ventilator pressure data [7] [10].

Let *P*_*t*_ denote the pressure at time *t*, and let *P*_*t*−1_, *P*_*t*−2_, …, *P*_*t*−*d*_ represent the historical pressure data over the look-back period *d*. The first LSTM layer takes this pressure data as input and outputs a sequence of hidden state vectors 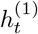:

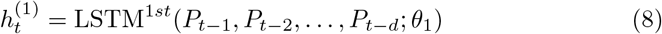

where *θ*_1_ are the parameters of the first LSTM layer. Each vector in 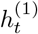 captures temporal dependencies, with dimensionality *N*_1_ *× d*, where *N*_1_ is the number of neurons in the first layer.

This sequence 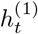 is then processed by a second LSTM layer to generate a single temporal feature vector 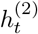:

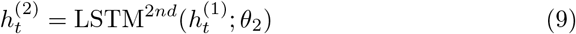

The output 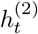, with dimensionality *N*_2_ *×* 1, encapsulates the complete temporal dependencies and serves as input to DNN1.

While LSTM effectively captures temporal aspects, it may not fully address non-linear interactions. DNN1 addresses this by processing the LSTM output to extract higher-order relationships, defined as:

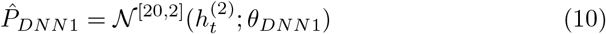

Simultaneously, DNN2 extracts non-linear relationships from a distinct feature vector *X* ∈ ℝ^6,036,000*×*7^:

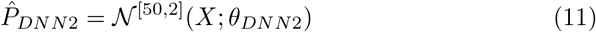

The outputs of DNN1 and DNN2 are concatenated and passed through a final output layer:

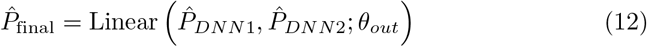

This hybrid architecture effectively combines the strengths of both networks, enhancing the model’s ability to predict ventilator pressure accurately, as illustrated in Figure 8.

**Fig 8**. The structure of hybrid model [Figure12]

### Forget Gate Reset Mechanism

A critical feature of our hybrid LSTM-DNN model is the reset of the forget gate, which preserves prediction integrity across breath cycles. After processing each breath cycle of 80 time steps, the hidden state *h*_*t*_ and cell state *c*_*t*_ are reset to zero:

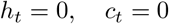

This ensures the model captures unique patterns for each cycle without interference from prior data, enhancing prediction accuracy [32].

### the Experimental Setup

The configuration for the hybrid LSTM-DNN model experiment focuses on predicting ventilator pressure using a well-defined architecture that integrates two LSTM layers and multiple DNN components. The model is configured with a time step of four, utilizing a single feature for the LSTM input, while the DNN processes a total of seven features. An initial learning rate of 0.001 is employed, which is adjusted through an exponential decay schedule to enhance training stability. With a batch size of 256, the model ensures efficient data processing during training. Dropout regularization is applied at a rate of 40% after the first dense layer in each DNN network and also at the output layer to mitigate overfitting.

## Evaluation Techniques

### Overview of Data-Splitting Methods

Cross-validation is a key data resampling technique in machine learning, essential for evaluating model generalization and minimizing overfitting [12]. As part of the Monte Carlo methods, it provides robust performance estimates [28]. K-fold cross-validation enhances reliability by partitioning data into *K* subsets [8].

Let *D* represent our dataset, consisting of *N* = 6, 036, 000 samples, denoted by *D* = {*x*_1_, *x*_2_, …, *x*_*N*_}, where each *x*_*i*_ is a feature vector. Under K-fold cross-validation, the dataset *D* is partitioned into *K* equally sized (or nearly equal) subsets, represented as *D*_1_, *D*_2_, …, *D*_*K*_.

The size of each fold can be calculated using the formula [8]:

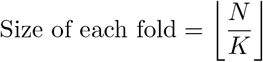

For each fold *k* (where *k* = 1, 2, …, *K*):

- We designate *D*_test_ = *D*_*k*_ as the validation set, while the training set is defined as *D*_train_ = _*j*≠*k*_ *D*_*j*_.
- Our hybrid model *h* is then trained on the training set *D*_train_ and subsequently evaluated on the validation set *D*_test_.

The training phase aims to determine the optimal values for the model parameters *θ* by minimizing the discrepancy between the predicted outputs and the true target values [1]. This optimization process fine-tunes *θ* to improve the model’s predictive accuracy, this means

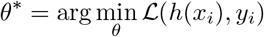

where ℒ represents the loss function that measures the difference between the predicted outputs *h*(*x*_*i*_) and the true values *y*_*i*_. The specifics of the evaluation metrics employed in this analysis will be detailed in the Evaluation Metrics section.

Upon completion of the training on *K* − 1 folds, predictions are made on the validation fold *D*_test_. Let 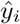 denote the predicted output for the samples in *D*_test_. The model’s performance for this fold can be quantified using a performance metric [30].

After repeating this process for all *K* folds, we obtain *K* performance scores, denoted as ℒ_1_, ℒ_2_, …, ℒ_*K*_. The overall performance of the model is determined by averaging these scores:

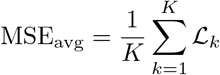

This systematic approach enables us to effectively evaluate the generalization capabilities of our hybrid model. However, it is important to note that while K-fold cross-validation is beneficial for static datasets, it is ineffective for time series data due to its disregard for temporal dependencies. To address this limitation, we employ time series splitting methods, which preserve the order of observations and prevent data leakage, ensuring accurate model evaluation.

Let *Y* = *{Y*_1_, *Y*_2_, …, *Y*_*T*_ *}* represent our time series dataset (pressure), where each *Y*_*i*_ denotes the observation at time step *i*. Each observation *Y*_*i*_ can be defined as a vector of values over a specified step size *s*:

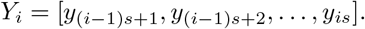

To facilitate an effective training and validation process, we introduce a gap parameter *g*, which indicates the number of time steps to be skipped between the end of the training set and the beginning of the validation set. This gap ensures that the validation data remains temporally distinct from the training data. For each fold *k* (where *k* = 1, 2, …, *K*), the training set is defined as

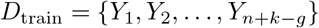

and the validation set as

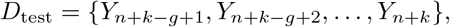

where *n* represents the size of each training window. The training process follows the standard K-fold methodology while maintaining the integrity of the time series structure.

### Evaluation Metrics

Evaluating predictive models is essential for assessing their real-world effectiveness. This study employs several metrics for our hybrid LSTM-DNN model in ventilator pressure prediction.

**Mean Absolute Error (MAE)** [16] [22] **measures the average absolute difference:**

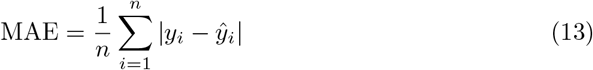

**Mean Squared Error (MSE)** [16] quantifies the average of the squared differences between predicted and actual values, highlighting larger errors:

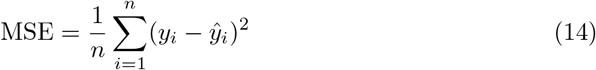

**Root Mean Squared Error (RMSE)** [18] [22] **measures the square root of the MSE:**

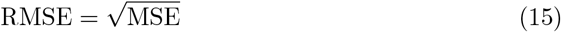

**Coefficient of Determination** (*R*^2^) [16] represents the percentage of variation in the dependent variable that is accounted for by the model:

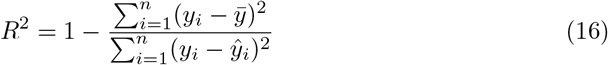

A higher *R*^2^ value (closer to 1) suggests a better model fit.

**Explained Variance Score (EVS)** [16] measures the proportion of variance in the dependent variable that can be predicted:

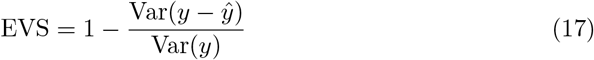

Higher EVS values (closer to 1) indicate better model performance.

**Experimental Results**

This section presents a detailed evaluation of the hybrid LSTM-DNN model’s performance, utilizing K-fold cross-validation and time-series split methods. We progressively exclude configurations identified as error-prone during each metric analysis to ensure clarity and accuracy in subsequent evaluations. This approach enables us to compare the effectiveness of different configurations and assess the impact of the LSTM forget gate reset mechanism on model accuracy. The section concludes with a benchmarking analysis against recent studies, highlighting the innovations and advantages of our model.

The K-fold cross-validation results, as seen in Table 3, show that minimizing *K* enhances the hybrid LSTM-DNN model’s robustness in predicting ventilator pressure.

**Table 3.** K-fold Cross-Validation Results.

| K | Train Loss | Val Loss | RMSE | MAE | EVS | R<sup>2</sup> |
| --- | --- | --- | --- | --- | --- | --- |
| 19 | 0.0028 | 0.0015 | 0.0525 | 0.0328 | 0.8153 | 0.8136 |
| 10 | 0.0025 | 0.0016 | 0.0501 | 0.0313 | 0.8318 | 0.8304 |
| 7 | 0.0023 | 0.0013 | 0.0484 | 0.0302 | 0.8420 | 0.8409 |
| 3 | 0.0019 | 0.0009 | 0.0439 | 0.0269 | 0.8697 | 0.8689 |

Building on the K-fold cross-validation results, we assess our model with time series split to evaluate temporal dependencies.

**Fig 9**. Training loss for different configurations [Figure13]

Figure 9 shows that configurations *K_*10*_gap_*0 and *K_*10*_gap_*1 are unreliable outliers, warranting their exclusion. In contrast, *K_*3*_gap_*1 and *K_*3*_gap_*10 achieve the lowest maximum training loss of 0.0022, demonstrating superior stability. Notably, configuration *K_*3*_gap_*0 has an interquartile range (IQR) of zero, indicating minimal variability in model performance. In contrast, configurations *K_*10*_gap_*10 and *K_*7*_gap_*1 exhibit the highest IQRs of 0.0005 and 0.000475, respectively, suggesting higher variability across these configurations.

**Fig 10**. Validation loss for different configurations [Figure14]

The validation loss analysis in Figure 10 highlights key insights into model stability. Configurations *K_*19*_gap_*1 and *K_*19*_gap_*10 are outliers and should be excluded due to distorted performance. Additionally, *K_*3*_gap_*1 has the highest IQR of 0.0024, indicating significant variability, justifying its removal. In contrast, *K_*3*_gap_*10 shows the lowest IQR of 0.00005, reflecting excellent stability. This evaluation emphasizes the need to refine the dataset for more reliable performance insights.

**Fig 11**. RMSE for different configurations [Figure15]

Figure 11 illustrates RMSE across configurations, showing that *K_*3*_gap_*10 achieves the lowest maximum RMSE of 0.0473, indicating superior performance. The smallest IQR of 0.00045 for *K_*3*_gap_*0 reflects low variability, enhancing its reliability. In contrast, *K_*7*_gap_*10 has the highest IQR of 0.004275, suggesting significant performance variability. This analysis sets the stage for further exploration of these results’ implications on overall model performance and stability.

**Fig 12**. MAE for different configurations [Figure16]

Figure 12 reveals that IQR for the configuration *K_*3*_gap_*0 is the smallest, measured at 0.0005, while *K_*3*_gap_*10 follows with an IQR of 0.00085.This indicates that *K_*3*_gap_*0 demonstrates lower variability in performance compared to *K_*3 *gap_*10, suggesting a potential advantage in stability for the former configuration. This distinction in IQR values may have important implications for the reliability of model assessments across these configurations.

**Fig 13**. EVS for different configurations [Figure17]

**Fig 14**. *R*^2^ for different configurations [Figure18]

According to Figures 13,14, the configuration *K_*3*_gap_*10 emerges as the most effective, achieving EVS of 0.8496 and an *R*^2^ value of 0.84865. These metrics highlight its exceptional performance compared to other configurations, reinforcing its reliability for time series cross-validation.

We evaluated the LSTM forget gate reset mechanism on the *K_*3*_gap_*10 configuration. Figures 15 and 16 illustrate improved training and validation losses, indicating enhanced convergence and reduced overfitting with the reset mechanism.

**(a)** Explained Variance [Figure19] **(b)** *R*^2^ [Figure20]

**Fig 15**. Comparison of *R*^2^ and Explained Variance with/without Reset Mechanism [Figure19, Figure20]

**Fig 16**. Comparison of error metrics with/without Reset Mechanism [Figure21]

The comparison in Table 4 illustrates the superior performance of our proposed Hybrid LSTM-DNN model, which achieves significantly lower error metrics in both RMSE (0.002) and MAE (0.02795) compared to previous models, demonstrating our model’s enhanced accuracy and predictive power in ventilator pressure forecasting.

**Table 4.**
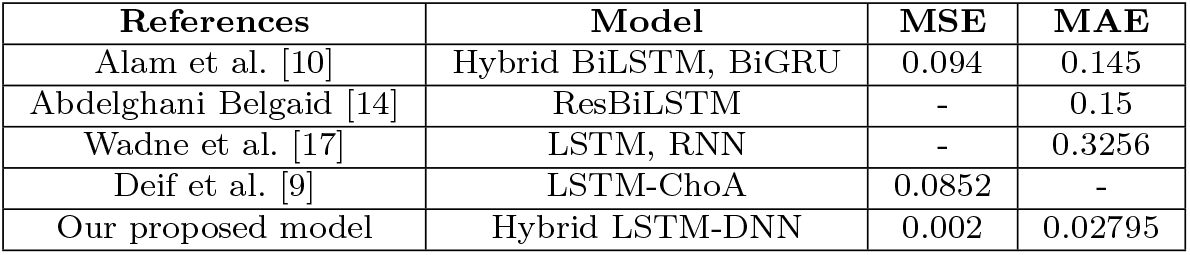
Comparing results of the proposed model with comparative studies.

## Discussion

Training confirms the model’s strong predictive performance, with the forget gate reset preventing data leakage and TimeSeriesSplit optimizing temporal continuity for clinical reliability. This enhances patient safety through accurate ventilator adjustments. Future adaptations could apply the model to other clinical time-series data, supporting real-time monitoring in critical care applications.

## Conclusion

This study introduced a hybrid LSTM-DNN model to improve ventilator pressure prediction by capturing temporal dependencies and non-linear relationships. A unique forget gate reset mechanism prevents data leakage, enhancing patient-specific accuracy. Evaluations with K-fold and TimeSeriesSplit showed robust performance, with TimeSeriesSplit better preserving temporal continuity. Optimized hyperparameters further stabilized training, underscoring the model’s suitability for clinical applications. Future work will explore refinements for broader medical forecasting.

## Data Availability

The data underlying the findings described in this manuscript are fully available without restriction. The dataset used in this study is publicly accessible on Kaggle at the following URL: [https://www.kaggle.com/competitions/ventilator-pressure-prediction/data]. The ventilator data, produced using a modified open-source ventilator connected to an artificial bellows test lung via a respiratory circuit, contains control inputs and the state variable (airway pressure) to predict. This dataset was utilized to train and evaluate the proposed Hybrid LSTM-DNN model for ventilator pressure prediction.

## References

1. Koca M, Avci I. A novel hybrid model detection of security vulnerabilities in industrial control systems and IoT using GCN+LSTM. IEEE Access. vol. 12, pp. 2024, 2024. Available from: 10.1109/ACCESS.2024.3466391

2. Al-Selwi SM, Hassan MF, Abdulkadir SJ, Muneer A, Sumiea EH, Alqushaibi A, Ragab MG. RNN-LSTM: From applications to modeling techniques and beyond—Systematic review. Journal of King Saud University - Computer and Information Sciences. 2024

3. Mahmoud A, Mohammed A. Leveraging hybrid deep learning models for enhanced multivariate time series forecasting. Neural Processing Letters. 2024;56:223. Available from: 10.1007/s11063-024-11656-3

4. Hickey SM, Sankari A, Giwa AO. Mechanical ventilation. National Center for Biotechnology Information. 2024. Available from: https://www.ncbi.nlm.nih.gov/books/NBK539742/

5. Gong Y, Sankari A. Noninvasive ventilation. In: StatPearls. StatPearls Publishing; 2024. Available from: https://www.ncbi.nlm.nih.gov/books/NBK578188/

6. Mortezapour Shiri F, Perumal T, Mustapha N, Mohamed R. A comprehensive overview and comparative analysis on deep learning models: CNN, RNN, LSTM, GRU. arXiv. 2023. Available from: 10.13140/RG.2.2.11938.81609

7. Al-Ghamdi M, Al-Ghamdi AA, Ragab M. A hybrid DNN multilayered LSTM model for energy consumption prediction. Applied Sciences. 2023;13(20):11408. Available from: 10.3390/app132011408

8. Oyedele O. Determining the optimal number of folds to use in a K-fold cross-validation: A neural network classification experiment. Research in Mathematics. 2023;10(1):2201015. Available from: 10.1080/27684830.2023.2201015

9. Ahmed FR, Alsenany SA, Abdelaliem SMF, Deif MA. Development of a hybrid LSTM with chimp optimization algorithm for the pressure ventilator prediction. Scientific Reports. 2023;13:20927. Available from: 10.1038/s41598-023-47837-8

10. Alam MJ, Rabbi J, Ahamed S. Forecasting pressure of ventilator using a hybrid deep learning model built with Bi-LSTM and Bi-GRU to simulate ventilation. arXiv. 2023. Available from: https://arxiv.org/abs/2302.09691/

11. Ozcan A, Catal C, Donmez E, Senturk B. A hybrid DNN–LSTM model for detecting phishing URLs. Neural Computing and Applications. 2023;35:4957–4973. Available from: 10.1007/s00521-021-06401-z

12. Wilimitis D, Walsh CG. Practical considerations and applied examples of cross-validation for model development and evaluation in health care: Tutorial. JMIR AI. 2023;2:e49023. Available from: 10.2196/49023

13. Ibrahim RT, Mohamed YA, Abd El-kader MS, Azouz AM. Airway pressure release ventilation versus pressure-controlled ventilation in acute hypoxemic respiratory failure. The Egyptian Journal of Chest Diseases and Tuberculosis. 2022;71(1):74–80. Available from: 10.4103/ejcdt.ejcdt_82_20

14. Belgaid A. Deep sequence modeling for pressure controlled mechanical ventilation. medRxiv. 2022. Available from: 10.1101/2022.03.02.22271790

15. Tamal MBA, Alam MA, Sharker MN, Sazib MI. Forecasting of solar photovoltaic output energy using LSTM machine learning algorithm. In: 2022 4th International Conference on Sustainable Technologies for Industry 4.0 (STI). IEEE; 2022. p. 1–6. Available from: 10.1109/STI56238.2022.10103310

16. Plevris V, Solorzano G, Bakas NP, Ben Seghier MEA. Investigation of performance metrics in regression analysis and machine learning-based prediction models. In: Proceedings of the ECCOMAS Congress 2022.2022. Available from: 10.23967/eccomas.2022.155

17. Wadne V, Wanve O, Singh A, Hanabar Y, Wable S. Pressure prediction system in lung circuit using deep learning and machine learning. International Research Journal of Engineering and Technology (IRJET). 2022;9(5):3041.

18. Hodson TO. Root-mean-square error (RMSE) or mean absolute error (MAE): When to use them or not. Geoscientific Model Development. 2022;15(14):5481–5487. Available from: 10.5194/gmd-15-5481-2022

19. Hamdan MH, Roach D. The sigmoid neural network activation function and its connections to Airy’s and the Nield-Kuznetsov functions. University of New Brunswick. 2022. Available from: 10.37394/232020.2022.2.13

20. Lindemann B, Müller T, Vietz H, Jazdi N, Weyrich M. A survey on long short-term memory networks for time series prediction. In: Proceedings of the 14th CIRP Conference on Intelligent Computation in Manufacturing Engineering (CIRP ICME ‘20). 2021. p. 2212–8271. Available from: 10.1016/j.procir.2021.03.088

21. Sullivan J. Engineering and artificial intelligence combine to safeguard patients’ lives. Princeton University School of Engineering and Applied Science. 2021 Jan Available from: https://engineering.princeton.edu/news/2021/01/26/engineering-and-artificial-intelligence-combine-safeguard-patients-lives

22. Chicco D, Warrens MJ, Jurman G. The coefficient of determination R-squared is more informative than SMAPE, MAE, MAPE, MSE, and RMSE in regression analysis evaluation. PeerJ Computer Science. 2021;7(3):e623. Available from: 10.7717/peerj-cs.623

23. Van Houdt G, Mosquera C, Nápoles G. A review on the long short-term memory model. Artificial Intelligence Review. 2020;53(1). Available from: 10.1007/s10462-020-09838-1

24. Guèrin C, Lèvy P. Easier access to mechanical ventilation worldwide: An urgent need for low-income countries, especially in face of the growing COVID-19 crisis. European Respiratory Journal. 2020;55(6). Available from: 10.1183/13993003.01271-2020

25. Galván E, Mooney P. Neuroevolution in deep neural networks: Current trends and future challenges. arXiv preprint arXiv:2006.05415v1. 2020. Available from: https://arxiv.org/abs/2006.05415

26. de Haro C, Ochagavia A, López-Aguilar J, Fernandez-Gonzalo S, Navarra-Ventura G, Magrans R, Montanyà J, Blanch L, the ASYNICU Group. Patient-ventilator asynchronies during mechanical ventilation: Current knowledge and research priorities. Intensive Care Medicine Experimental. 2019;7(Suppl 1):43.

27. Ali MA, Zhuang H, Ibrahim A, Rehman O, Huang M, Wu A. A machine learning approach for the classification of kidney cancer subtypes using miRNA. Applied Sciences. 2018;8(12):2422. Available from: 10.3390/app8122422

28. Berrar D. Cross-validation. In: Encyclopedia of Bioinformatics and Computational Biology. Elsevier; 018. p. 542–545. Available from: 10.1016/B978-0-12-809633-8.20349-X

29. Botchkarev A. Evaluating performance of regression machine learning models using multiple error metrics in Azure Machine Learning Studio. SSRN Electronic Journal. 2018. Available from: 10.2139/ssrn.3177507

30. Ahmed FYH, Ali YH, Shamsuddin SM. Using K-fold cross-validation proposed models for Spikeprop learning enhancements. International Journal of Engineering & Technology. 2018;7(4.11):145–151.

31. Becker M. A deep neural network for robust feature extraction. Master’s thesis, University of Applied Sciences Düsseldorf, Faculty of Electrical Engineering & Information Technology. 2017. Available from: 10.13140/RG.2.2.35523.14887

32. Gulli A, Pal S. Deep Learning with Keras. Packt Publishing; 2017. ISBN 978-1-78712-842-2.

33. Paramasivam E, Bodenham A. Air leaks, pneumothorax, and chest drains. Continuing Education in Anaesthesia Critical Care & Pain. 2008;8(6):204–209. Available from: 10.1093/bjaceaccp/mkn038

